# An intratumorally discovered extracellular vesicle-derived biomarker enables ultrasensitive digital quantification in plasma for early detection of non-small cell lung cancer

**DOI:** 10.1101/2025.06.09.25329294

**Authors:** Heng Huang, Taketo Kato, Yuichi Abe, Akira Yokoi, Masami Kitagawa, Eri Asano-Inami, Yoshito Imamura, Yuji Nomata, Hiroki Watanabe, Hirofumi Takenaka, Taiki Ryo, Yuta Kawasumi, Keita Nakanishi, Yuka Kadomatsu, Harushi Ueno, Shota Nakamura, Tetsuya Mizuno, Ayumu Taguchi, Toyofumi Fengshi Chen-Yoshikawa

**Author notes:** These authors contribute to the work equally (Heng Huang, Taketo Kato). **Corresponding author:** Toyofumi Fengshi Chen-Yoshikawa, Department of Thoracic Surgery, Nagoya University Graduate School of Medicine, 65 Tsurumai-cho, Showa-ku, Nagoya City, Aichi, 466-8550, Japan.

## Abstract

**Background:** Extracellular vesicle (EV)-based liquid biopsy remains challenging due to the limited specificity and low abundance of EV cargoes. This study aimed to identify cancer-specific molecules from tissue-derived EVs and evaluate their potential as circulating biomarkers for non-small cell lung cancer (NSCLC).

**Methods:** Tissue-derived EVs were isolated by density gradient flotation and subjected to proteomic profiling to identify differentially expressed proteins. Public transcriptomic data, functional enrichment, and variable importance scores were integrated to select candidate biomarkers. Immunogold labeling transmission electron microscopy (TEM) was used to visualize protein expression on the EV membrane using, and an ultrasensitive modified digital EV screening technique (DEST) was employed to quantify protein signals in circulating EVs. The diagnostic performance was evaluated by the area under the curves (AUC).

**Results:** Tissue-derived EVs were isolated from cancer tissues and paired non-cancer lung tissues from patients with NSCLC. Proteomic profiling identified 1,568 tissue-derived EV proteins, of which 904 were differentially expressed. Integrated transcriptomic and proteomic analyses yielded eight cancer-specific, membrane-localized candidate biomarkers, with GLUT1 showing the highest variable importance score. Immunogold labeling TEM confirmed the localization of GLUT1 on the EV membrane. Modified DEST analysis of 47 patients revealed significantly elevated levels of CD63^+^ EV-derived GLUT1 in the cancer group than in the non-cancer group (*P* < 0.01). Diagnostic performance reached an AUC of 0.817 in the whole cohort and 0.898 in the stage-I cohort, exceeding that of conventional serum biomarkers. A three-marker panel (CD63^+^ EV-derived GLUT1, CYFRA 21-1, and CEA) further improved discrimination, with AUCs of 0.853 in the whole cohort and 0.944 in the stage-I cohort.

**Conclusion:** Circulating EV-derived GLUT1 was preliminarily identified as a novel biomarker for NSCLC, and its combination with conventional serum biomarkers enhanced diagnostic performance, particularly in early-stage cases.

## Introduction

Extracellular vesicles (EVs), present in diverse biological fluids, are increasingly recognized as essential mediators of intercellular communication. They contribute to alterations in both local tissues and systemic environments, promoting cancer cell proliferation and dissemination [1, 2]. These nanoscale vesicles, particularly exosomes, transport biomolecules such as proteins, DNA, and mRNAs that reflect the state of their cells of origin [3, 4]. Theoretically, cancer cell-derived EVs released into the circulation may harbor disease-specific molecules, providing promising biomarkers for minimally invasive liquid biopsy (LB) [5, 6]. Among the bioactive EV cargoes, membrane-localized proteins have been the most promising candidates for cancer diagnosis and EV-subset identification due to their accessibility, cellular specificity, and roles in EV targeting and uptake [7–9].

Non-small cell lung cancer (NSCLC) is the predominant subtype of lung cancer and remains the leading cause of cancer-related death worldwide [10, 11]. In addition to suboptimal therapeutic strategies, delayed detection has significantly contributed to poor clinical outcomes in patients with NSCLC [12, 13]. Contemporary diagnostic paradigms for NSCLC encompass tissue biopsy, imaging examinations, and serum biomarkers, but these methods are limited by invasiveness, their intratumoral heterogeneity, and unsatisfactory specificity or sensitivity [14–16]. The development of novel EV-based biomarkers with superior diagnostic performance is a widely discussed and prioritized topic, offering new opportunities for early detection and improved management of NSCLC [6]. Importantly, alterations in EV cargoes may precede detectable changes in conventional serum biomarkers or imaging findings [17, 18].

However, none of the identified circulating EV-derived biomarkers have been effectively translated into clinical practice [19]. In particular, conventional tools often fail to detect the scarce signals from low-abundance circulating EV proteins [20, 21]. Therefore, it is critical to integrate robust signal amplification strategies with high-specificity targeting molecules to develop accurate and sensitive diagnostic systems [22]. Tyramide signal amplification (TSA) is one such method in which the detection antibody conjugated to horseradish peroxidase (HRP) catalyzes the conversion of biotinylated tyramide into highly reactive radicals. These activated radicals covalently bind to the surrounding tyrosine residues, resulting in the localized deposition of stable biotin molecules at the HRP-binding sites and substantial amplification of the target signals [18, 23]. This approach is increasingly applied in various immunoassays of EV-related research [22, 24–27].

The aim of this study was to integrate tissue-derived EV proteomic profiling with public transcriptomic data to identify novel biomarkers for NSCLC. This effort led to the selection of glucose transporter 1 (GLUT1) as a promising EV-derived and membrane-localized biomarker candidate. We then verified the favorable diagnostic performance of circulating EV-derived GLUT1 in clinical NSCLC, particularly in early-stage cases. To overcome technical challenges in detecting scarce signals, we applied an ultrasensitive modified digital EV screening technique (DEST) incorporating a TSA step, which enabled rapid and sensitive detection of low-abundance EV-derived proteins from minimally processed plasma samples [26].

## Materials and methods

### 1. Clinical sample preparation

The current study was conducted in accordance with the ethical guidelines and received approval from the Institutional Review Board of Nagoya University Hospital (No.2022-0107). The tissue and peripheral blood specimens were collected at Nagoya University Hospital between 2022 and 2023. None of the patients had received preoperative anticancer therapy, or had any co-existing malignancies. Whole blood was drawn into four purple-top EDTA tubes, kept upright at 4 ℃, and processed within 1 hour of collection. After plasma separation, the samples were transferred to 15 ml tubes and centrifuged at 1500 × g for 10 minutes at 4 ℃. The resulting supernatant was aliquoted and stored at -80 °C until further use.

### 2. Maintenance of cell lines

The HCC827 and A549 cell lines were kindly provided by the Department of Respiratory Medicine, Nagoya University Hospital, and were cultured in Roswell Park Memorial Institute (RPMI) 1640 medium (Thermo Fisher Scientific) supplemented with 10% fetal bovine serum (FBS; Gibco) and 1% penicillin-streptomycin (PS; Gibco).

### 3. Isolation of EVs by density gradient flotation

Tissue-derived EVs were isolated by ultracentrifugation with density gradient flotation [28]. Tissue specimens were digested enzymatically for 1 hour at 37 ℃ using the Tumor Dissociation Kit (Miltenyi Biotec) in a gentleMACS Octo Dissociator with Heaters. The resulting suspension was centrifuged at 3,000 × g for 15 minutes to remove tissue debris. The supernatant was transferred into Polypropylene Centrifuge Tubes (Beckman Coulter Inc., USA) and mixed with filtered iodixanol solution to a final density of ∼1.163-1.189 g/ml. A 2 ml layer of 1.134 g/ml iodixanol solution was carefully overlaid to establish a discontinuous density gradient. Ultracentrifugation was performed at 65,000 × g for more than 12 hours at 4°C using an Optima XE-100 ultracentrifuge with an SW 41 Ti Swinging-Bucket rotor (Beckman Coulter Inc., USA). A 500 μl fraction of the supernatant was collected and filtered through 0.22 μm membrane filters for subsequent characterization (**Figure 1A**).

**Figure 1.**
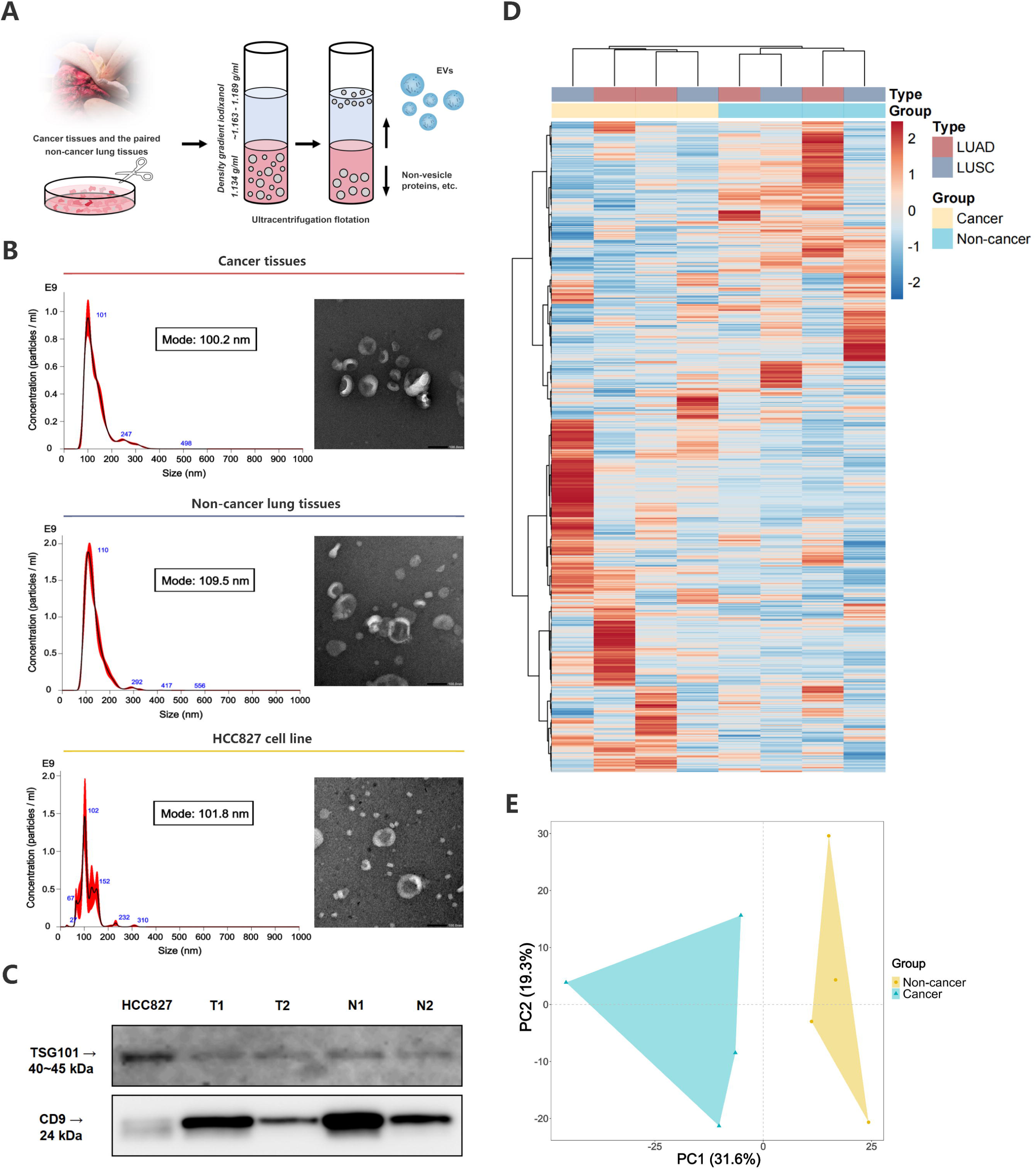
Isolation and characterization of tissue-derived EVs. (**A**) The schematic illustration of EV isolation. (**B**) NTA-based EV characterization, combined with TEM images, and (**C**) WB analysis of EV markers CD9 and TSG101 from representative tissue samples and the HCC827 cell line. (**D**) Hierarchical clustering heatmap illustrating proteomic profiles from tissue-derived EVs. (**E**) PCA plot of different tissue-derived EVs based on proteomic profiles. **Abbreviations**: EV, extracellular vesicle; LUAD, lung adenocarcinoma; LUSC, lung squamous carcinoma; NTA, nanoparticle tracking analysis; TEM, transmission electron microscopy; WB, western blotting; PCA, principal component analysis

For the isolation of cell-derived EVs, the culture medium was replaced with Advanced RPMI 1640 medium (Thermo Fisher Scientific) supplemented with 1% EV-depleted FBS at ∼70% cell confluence. Then the conditioned media were collected and centrifuged at 2,000 × g for 10 minutes to remove cell debris, followed by filtration through a Vacuum Filter/Storage Bottle System (150 ml, 0.22 μm diameter; Corning). The filtered medium was then aliquoted into Amicon^®^ Ultra-15ml Centrifugal Filters and centrifuged at ∼2,600 × g. Ultracentrifugation was performed using the same procedure as for tissue-derived EVs. After that, 1 ml of supernatant was transferred to Amicon^®^ Ultra-2ml Centrifugal Filters, and centrifuged at ∼2,600 × g until the medium was concentrated to about 150 μl for subsequent characterization.

### 4. Characterization of EVs

#### 4.1 Nanoparticle tracking analysis (NTA)

Size distribution, particle concentration, and mode size of EVs were quantitatively analyzed using NTA with the NanoSight NS300 system (Malvern Panalytical Ltd., UK). EV samples were diluted 1:100 in phosphate-buffered saline (PBS). Triplicate measurements were performed for each sample, with individual recording duration of 60 seconds to ensure statistical reliability.

#### 4.2 Transmission electron microscopy (TEM)

For morphological assessment, 10 μl of EV suspension was loaded onto carbon-coated copper grids (Nisshin EM, Japan), and negatively stained with 1% aqueous uranyl acetate, followed by air dry at room temperature. For immunogold labeling, 5 μl of EV suspension was loaded onto carbon-coated grids and processed similarly, followed by blocking with 1% bovine serum albumin (BSA) for 1 hour at room temperature. Grids were then incubated with a primary antibody (Anti-Glucose Transporter GLUT1 antibody, Abcam, ab115730, rabbit, dilution 1:50) for 1 hour at room temperature. After PBS washes, grids were incubated with a 10-nm gold-labeled secondary antibody [anti-IgG (H+L) Rabbit Goat-Poly Gold 10nm, EMGAR 10, CRL, Cardiff, UK, dilution 1:50] for another hour. After PBS washes, the grids were fixed with 1% glutaraldehyde for 10 minutes, rinsed with distilled water, stained with 1% uranyl acetate, blotted with filter paper, and air-dried. Imaging was performed using a JEM-1400Plus TEM (JEOL Ltd., Japan).

#### 4.3 Western blotting (WB)

EV-derived protein quantification was performed using the Qubit™ Protein and Protein Broad Range Assay Kits (Thermo Fisher Scientific) with the Qubit 4.0 Fluorometer (Invitrogen Co., USA). EV samples were subjected to precast polyacrylamide gel electrophoresis at a constant current of 42 mA (ATTO Corporation). Then proteins were transferred onto polyvinylidene difluoride (PVDF) membranes at 24 V for 15 minutes using a semi-dry transfer system (ATTO Corporation). Membranes were blocked with 5% (w/v) skim milk (MORINAGA & CO., LTD., Japan) in Tris-buffered saline with Tween 20 (TBST, ATTO Corporation) for 1 hour at room temperature, followed by 3 TBST washes and overnight incubation at 4°C with primary antibodies: rabbit monoclonal anti-TSG101 (Abcam, ab125011, dilution 1:500), rabbit monoclonal anti-CD9 (Cell Signaling Technology, D8O1A, dilution 1:1000). After 3 TBST washes, membranes were incubated for 2 hours at room temperature with HRP-linked goat anti-rabbit IgG secondary antibody (Cell Signaling Technology, 7074S; dilution 1:2000). After 3 TBST washes, protein bands were visualized using the ImageQuant LAS 4010 system (GE Healthcare, IL, USA).

### 5. Mass spectrometry (MS)-based proteomic profiling

EVs isolated from tissues of eight NSCLC patients, as well as cultured human lung cancer cell lines, were lysed in 1% sodium dodecyl sulfate (SDS; FUJIFILM Wako) supplemented with a cOmplete™ protease inhibitor cocktail (Roche), followed by boiling at 100 °C for 5 minutes and sonication at 4 °C for 15 minutes. Ten micrograms of total protein were subjected to reduction with dithiothreitol (FUJIFILM Wako) and alkylation with iodoacetamide (Nacalai). Proteins were precipitated onto carboxylate-modified magnetic particles (Sera-Mag SpeedBeads; Cytiva) in 50% ethanol (FUJIFILM Wako) and washed three times with 80% ethanol to remove contaminants. The resulting protein aggregates were resuspended in 50 mM ammonium bicarbonate (FUJIFILM Wako) containing trypsin (Roche) at a protein-to-enzyme ratio of 50:1 and digested overnight at 37 °C. Digestion was terminated by the addition of 1% trifluoroacetic acid (FUJIFILM Wako).

Peptides equivalent to 8 µg of protein were desalted using C18 StageTips. Desalted peptides were labeled with Tandem Mass Tag (TMT) 10-plex reagents (Thermo Fisher Scientific) according to the manufacturer’s instructions. A total of 24 µg of the TMT-labeled peptide mixture (corresponding to 80 µg of protein) was fractionated into seven fractions on a C18/SCX StageTip using via pH and salt-based elution, followed by vacuum evaporation.

Each fraction was analyzed on a Q Exactive mass spectrometer coupled with an UltiMate 3000 nanoLC system (Thermo Fisher Scientific, USA), equipped with a nano-electrospray ionization (nano-ESI) source (AMR, Japan) and a 150 mm × 75 µm inner diameter capillary column (Nikkyo Technos, Japan). Low-pH reverse-phase liquid chromatography (RPLC) was performed at a flow rate of 300 nL/min using a linear gradient elution starting from 5% to 40% solvent B (95% acetonitrile with 0.1% formic acid) over 100 minutes. Solvent A consisted of 2% acetonitrile with 0.1% formic acid. MS1 scans were acquired in the mass-to-charge (m/z) range of 400-1600. The top 20 most intense precursor ions were selected for higher-energy collisional dissociation (HCD) and MS/MS analysis in a data-dependent acquisition (DDA) mode.

Protein identification and quantification were performed using MaxQuant software (version 1.6.7.0; http://maxquant.org) with the integrated Andromeda search engine. MS/MS spectra were searched against the UniProt human reference proteome database (release 2019_04; https://www.uniprot.org/), supplemented with 262 common contaminant proteins. Trypsin/P was specified as the digestion enzyme, allowing up to two missed cleavages. Carbamidomethylation of cysteine residues and TMT labeling of lysine residues and peptide N-termini were set as fixed modifications. Oxidation of methionine and deamidation of asparagine and glutamine were considered variable modifications. The false discovery rate (FDR) was controlled to be < 1% at the levels of protein groups, peptide groups, and peptide-spectrum matches (PSMs). Quantitative values for each protein group were median-normalized and log2-transformed prior to statistical analysis. Differentially expressed proteins (DEPs) of tissue-derived EVs were identified using Student’s t-test based on predefined significance thresholds.

### 6. Transcriptomic data analysis from public database

Publicly available transcriptomic data and matched clinical information for NSCLC patients were obtained from The Cancer Genome Atlas (TCGA) via Genome Data General Database (GDC) data portal and analyzed using bioinformatic methods. A custom Wilcoxon test-based function was applied to identify differentially expressed genes (DEGs) between cancer and non-cancer tissues, using criteria of |log(fold change)| ≥ 1 and FDR < 0.05. A gene set encoding plasma membrane-localized proteins was retrieved from the Human Protein Atlas (HPA) database, and served as a reference to screen the candidate target DEGs.

### 7. Immunofluorescence for protein localization

HCC827 and A549 cells were seeded onto 4-well chamber slides (WATSON BIO LAB) and cultured to ∼70% confluence. Cells were washed twice with PBS, fixed with 10% formalin for 15 minutes, and then permeabilized with 0.1% Triton X-100 (Sigma) in PBS for 5 minutes, followed by two additional washes. Non-specific binding sites were blocked with 0.5% BSA for 30 minutes at room temperature. Samples were incubated overnight at 4°C with a primary antibody (Anti-Glucose Transporter GLUT1 antibody, Abcam, ab115730, rabbit, dilution 1:100) diluted in 0.5% BSA. After two PBS washes, cells were incubated with a secondary antibody (Goat Anti-Rabbit IgG H&L, Alexa Fluor 488-conjugated, Abcam, ab150077, dilution 1:500) for 1 hour at room temperature in the dark, followed by two washes. Slides were briefly air-dried, and nuclei were counterstained with one drop of DAPI Fluoromount-G (Cosmo Bio). Coverslips were mounted onto glass slides and sealed by nail polish. Fluorescence images were captured using a KEYENCE, BZ-9000 fluorescence microscope at 40× magnification. Merged images of DAPI (blue) and GLUT1 (green) channels were generated to assess the subcellular localization of GLUT1.

### 8. Modified DEST assay

Experimental protocols of the modified DEST assay, including reagent specifications, incubation time and concentrations, were outlined in **Table S1** and **Table S2**. Beads readouts were acquired using a BD FACS Canto™ II Flow Cytometer with the following settings: forward scatter (530 V), side scatter (390 V), and Pacific blue (530 V). A polygon gate was applied to distinguish positive from negative bead populations. All data are reported as the percentage of positive beads out of 10,000 total events. The effective signal was calculated as the difference in positive bead percentages between the detection group and the blank control group.

### 9. Statistical analysis

All statistical analyses and data visualizations were conducted using R software (R Foundation for Statistical Computing, v4.3.3). Continuous variables were assessed using Student’s t-test or Mann-Whitney U test, and presented as mean ± standard deviation (SD) or median (interquartile range [IQR]). Categorical variables were reported as percentages. Unsupervised hierarchical clustering heatmaps were generated using the heatmap package (v1.0.12). Principal component analysis (PCA) plots, volcano plots, and box plots were created using ggplot2 package (v3.5.1). Functional enrichment analyses were conducted using the clusterProfiler package (v4.10.1). Feature importance scores, representing the relative contribution of each variable to the model’s predictive performance, were calculated using the randomForest package (v4.7-1.2). Kaplan-Meier curves were generated with the online tool Kaplan-Meier Plotter (https://kmplot.com). Receiver operating characteristic (ROC) curve analyses and area under the curve (AUC) values were computed using the pROC package (v1.18.5). Calibration curves were plotted using the ggplot2 package (version 3.5.1). The Brier scores (computed through 1,000 bootstrap resampling iterations) quantified predictive accuracy as the mean squared difference between estimated probabilities and actual outcomes; lower scores indicate better performance. Statistical significance was defined as *P* < 0.05.

## Results

### 1. Isolation and proteomic profiling of tissue-derived EVs

EVs were isolated from cancer tissues and their paired non-cancer lung tissue. A schematic illustration of ultracentrifugation with density gradient flotation was presented in **Figure 1A**. The NTA confirmed effective EV enrichment, revealing characteristic size distributions with mode sizes of 100.2 nm for cancer tissue-derived EVs, 109.5 nm for non-cancer tissue-derived EVs, and 101.8 nm for cell-derived EVs (**Figure 1B**). The TEM visualized EVs with an appropriate size range and representative cup-shaped morphology (**Figure 1B**). The WB results further confirmed the enrichment of EV markers (TSG101 and CD9) in tissue and cell-derived EVs (**Figure 1C**).

Proteomic profiling of tissue-derived EVs identified 1,675 proteins. After excluding 84 unannotated proteins and 23 with missing values, 1,568 were retained for downstream analysis. The hierarchical clustering heatmap revealed distinct protein expression patterns between the cancer and non-cancer groups (**Figure 1D**). The PCA plot demonstrated that the top two principal components (PC1 and PC2) collectively explained 50.9% of the total variance, and clearly separating cancer and non-cancer samples. (**Figure 1E**).

### 2. Identification and functional annotation of DEPs

We applied relaxed criteria for DEPs identification, with an absolute fold change (FC) threshold > 1.2 or < 0.83, to retain biologically relevant low-abundance EV proteins while maintaining statistical robustness. This approach yielded a final set of 904 DEPs for subsequent candidate target selection (**NUH group**, **Figure 2A**). The volcano plot demonstrated 544 up-regulated and 360 down-regulated DEPs (**Figure 2B**). The heatmap further visualized the DEPs with distinct expression patterns between the cancer and non-cancer groups (**Figure 2C**). The PCA plot revealed that the top two PCs accounted for 61.2% of the total variance, with clear separation (**Figure 2D**).

**Figure 2.**
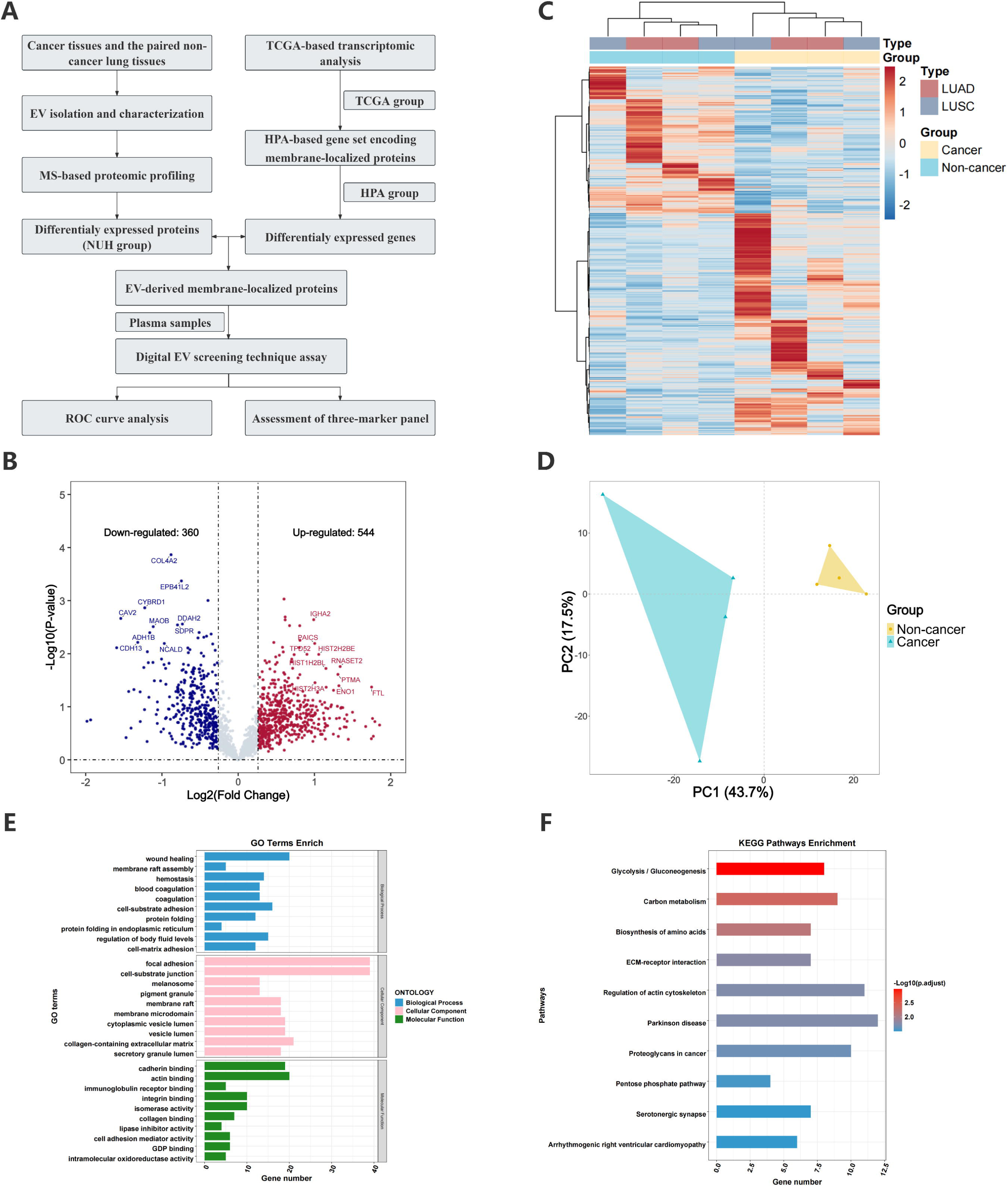
Identification and functional annotation of the DEPs from tissue-derived EVs. (**A**) The workflow for the discovery and clinical validation of circulating EV-derived biomarkers in NSCLC. (**B**) Volcano plot visualizing the distribution of DEPs. (**C**) Hierarchical clustering heatmap of DEPs from tissue-derived EVs. (**D**) PCA plot of different tissue-derived EVs based on DEPs. (**E**) GO enrichment analysis, and (**F**) KEGG pathway enrichment analysis of DEPs. **Abbreviations**: DEPs, differentially expressed proteins; EV, extracellular vesicle; NSCLC, non-small cell lung cancer; MS, mass spectrometry; NUH, Nagoya University Hospital; TCGA, The Cancer Genome Atlas; HPA, Human Protein Atlas; ROC, receiver operating characteristic; LUAD, lung adenocarcinoma; LUSC, lung squamous carcinoma; PCA, principal component analysis; GO, Gene Ontology; KEGG, Kyoto Encyclopedia of Genes and Genomes.

Functional annotations were performed on the top 200 up-regulated and 200 down-regulated DEPs (total N = 400) using Gene Ontology (GO) and Kyoto Encyclopedia of Genes and Genomes (KEGG) enrichment analyses. GO analysis indicated that DEPs were predominantly enriched in pathways related to adhesion, extracellular matrix remodeling, and hemostasis, suggesting their potential roles in tumor progression and EV-mediated modulation of the tumor microenvironment (**Figure 2E**). KEGG analysis identified several notable metabolism-related pathways, including glycolysis/gluconeogenesis, carbon metabolism, and the pentose phosphate pathway (**Figure 2F**).

### 3. Transcriptomic analysis and candidate target selection

Using TCGA transcriptomic data, 14,431 DEGs were identified from 973 cancer and 107 non-cancer samples, including 11,711 up-regulated and 2,720 down-regulated genes (**TCGA group**, **Figure S1A-B**). GO and KEGG enrichment analyses of these DEGs were presented in supplementary materials (**Figure S1C-D**). In parallel, a reference set of 2,074 genes encoding plasma membrane-localized proteins was curated from the HPA database (**HPA group**). Examination of the intersection of the NUH, TCGA, and HPA groups led to the identification of 24 DEGs encoding plasma membrane-localized proteins. Among these, 18 genes demonstrated consistent expression patterns in transcriptomic and proteomic analyses (**Figure 3A**). GO and KEGG analyses of these 18 DEGs predominantly involved glucose transport function, metabolism- and hypoxia-associated pathways (**Figure S1E-F**). Finally, eight genes with cancer-specific expression patterns were prioritized for further investigations, including SLC2A1, S100A2, ADH7, AKR1B10, IGKV3D-7, VIL1, FKBP2, and NME2 (**Figure 3A**).

**Figure 3.**
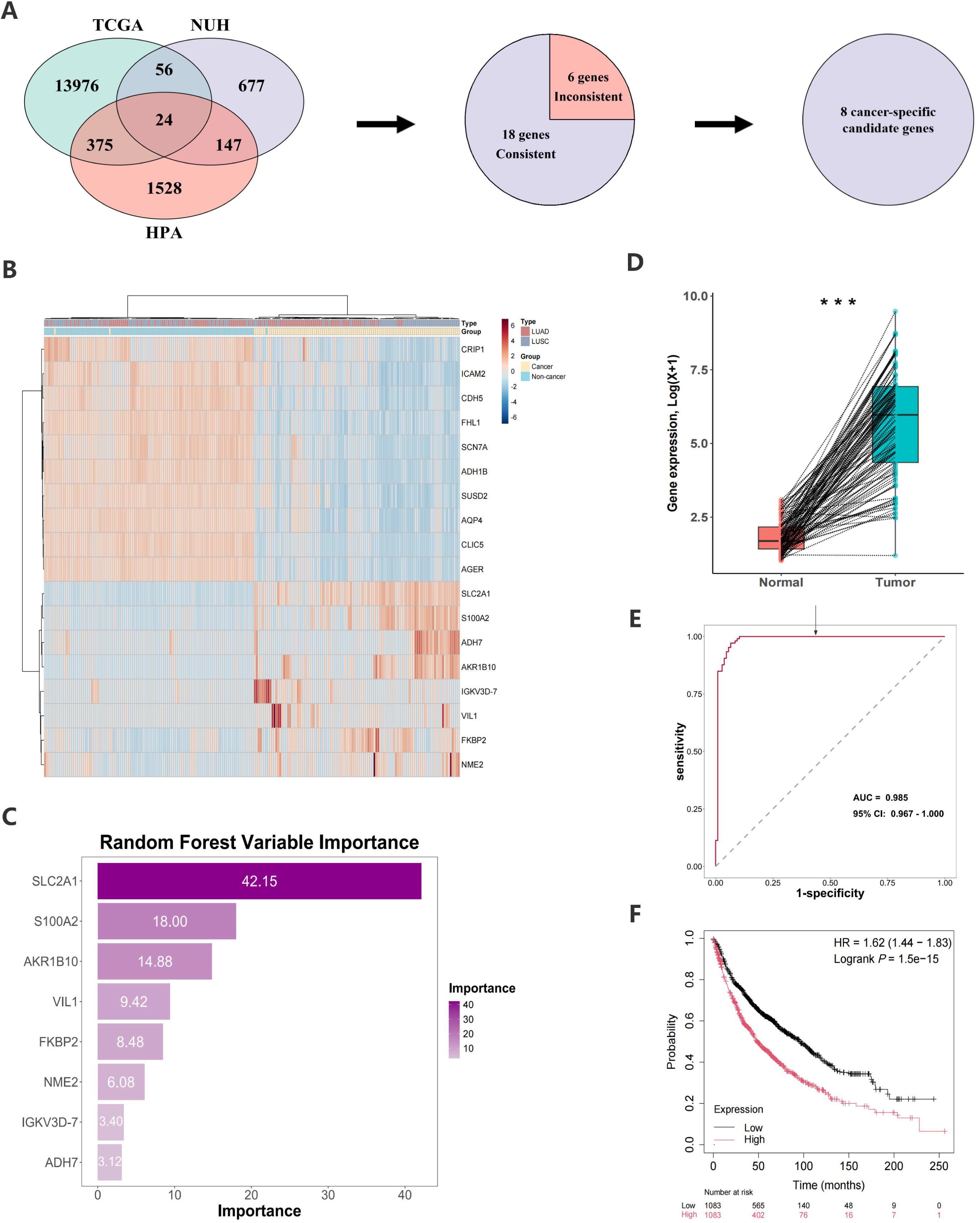
Selection of membrane-localized and cancer-specific candidate biomarkers. (**A**) Venn diagram illustrating the intersection of biomarkers across the three datasets. (**B**) Hierarchical clustering heatmap of 18 candidate target biomarkers from paired tissue samples. (**C**) Variable-importance bar plot ranking the cancer-specific candidate target biomarkers. (**D**) Box plot illustrating differential expression of SLC2A1. (**E**) ROC curve analysis evaluating the diagnostic performance of SLC2A1. (**F**) Kaplan-Meier curves demonstrating overall survival stratified by SLC2A1 expression. **Abbreviations**: TCGA, The Cancer Genome Atlas; HPA, Human Protein Atlas; NUH, Nagoya University Hospital; ROC, receiver operating characteristic; LUAD, lung adenocarcinoma; LUSC, lung squamous carcinoma; SLC2A1, solute carrier family 2 member 1. * All analyses in panels B-F were performed on 106 paired tumor-normal tissue samples from the TCGA dataset.

On the basis of the matched patient barcode, we created an internal validation cohort by selecting 106 paired cancer and non-cancer lung tissue samples from the TCGA NSCLC datasets. Distinctly different gene-expression patterns between the cancer and non-cancer samples were observed on the heatmap (**Figure 3B**). Eight candidate genes were ranked in variable importance analysis by a random-forest model constructed from these samples and SLC2A1 was assigned the highest score (**Figure 3C**). The SLC2A1 gene exhibited the most consistent and highest expression in cancer tissues among the other candidate genes (**Figure 3D, Figure S2A**). The ROC curve analysis also yielded the highest AUC value of 0.985 (95% CI: 0.967-1.000) (**Figure 3E, Figure S2B**). Kaplan-Meier survival analysis revealed significantly decreased overall survival in the patients with higher SLC2A1 expression (*P* < 0.001, **Figure 3F**)

### 4. Localization of GLUT1 (encoded by SLC2A1)

Although GLUT1 is a well-characterized cellular transmembrane protein, there is currently no evidence indicating its specific localization in EVs. To confirm the subcellular distribution of GLUT1, we performed immunofluorescence analysis in the HCC827 and A549 cell lines. As illustrated in **Figure 4A**, GLUT1 exhibited strong membrane-localized signals along the plasma membranes in both cell lines. In the EVs isolated from the conditioned media of these cell lines, immunogold labeling TEM clearly showed gold particles indicating GLUT1 localization on the vesicle surface (**Figure 4B**). These findings collectively demonstrate that GLUT1 is predominantly distributed on the membranes of cancer cells and their secreted EVs, indicating the technical feasibility for subsequent detection using the modified DEST assay.

**Figure 4.**
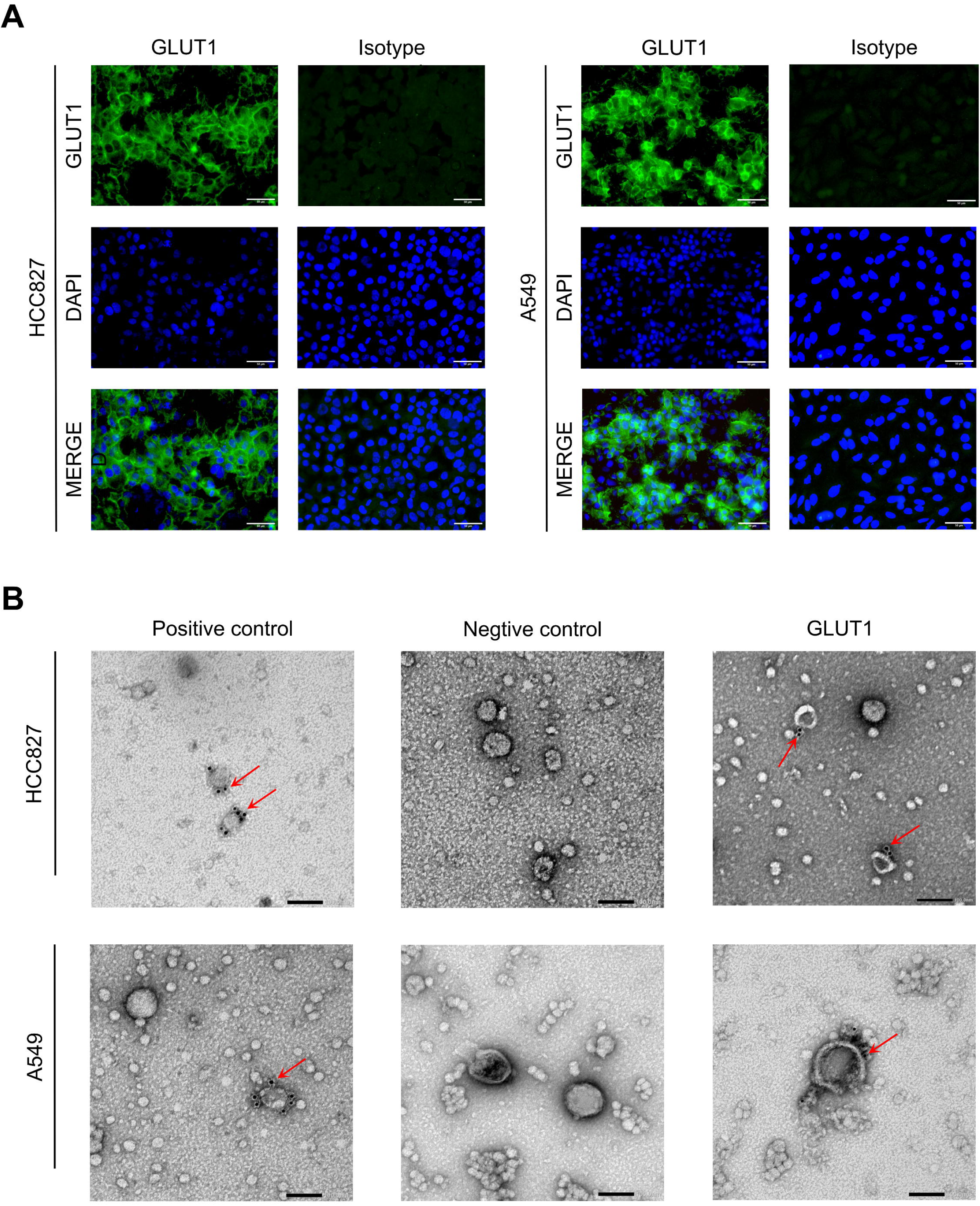
Subcellular and vesicular localization of GLUT1. (**A**) Immunofluorescence images of HCC827 and A549 cell lines stained for GLUT1 (green) with DAPI nuclear counter-stain (blue). (**B**) Immunogold labeling TEM of EVs derived from HCC827 and A549 cell lines. Red arrows indicate EVs positive for the target marker. **Abbreviations**: GLUT1, glucose transporter 1; EVs, extracellular vesicles; TEM, transmission electron microscopy.

### 5. Clinical validation based on the modified DEST assay

Plasma specimens from 38 histologically confirmed cancer patients and nine non-cancer controls were collected for use in the modified DEST assay to validate the circulating EV-derived GLUT1 expression in clinical NSCLC (**Table 1**). The schematic workflow and principle of this assay are presented in **Figure 5A**.

**Figure 5.**
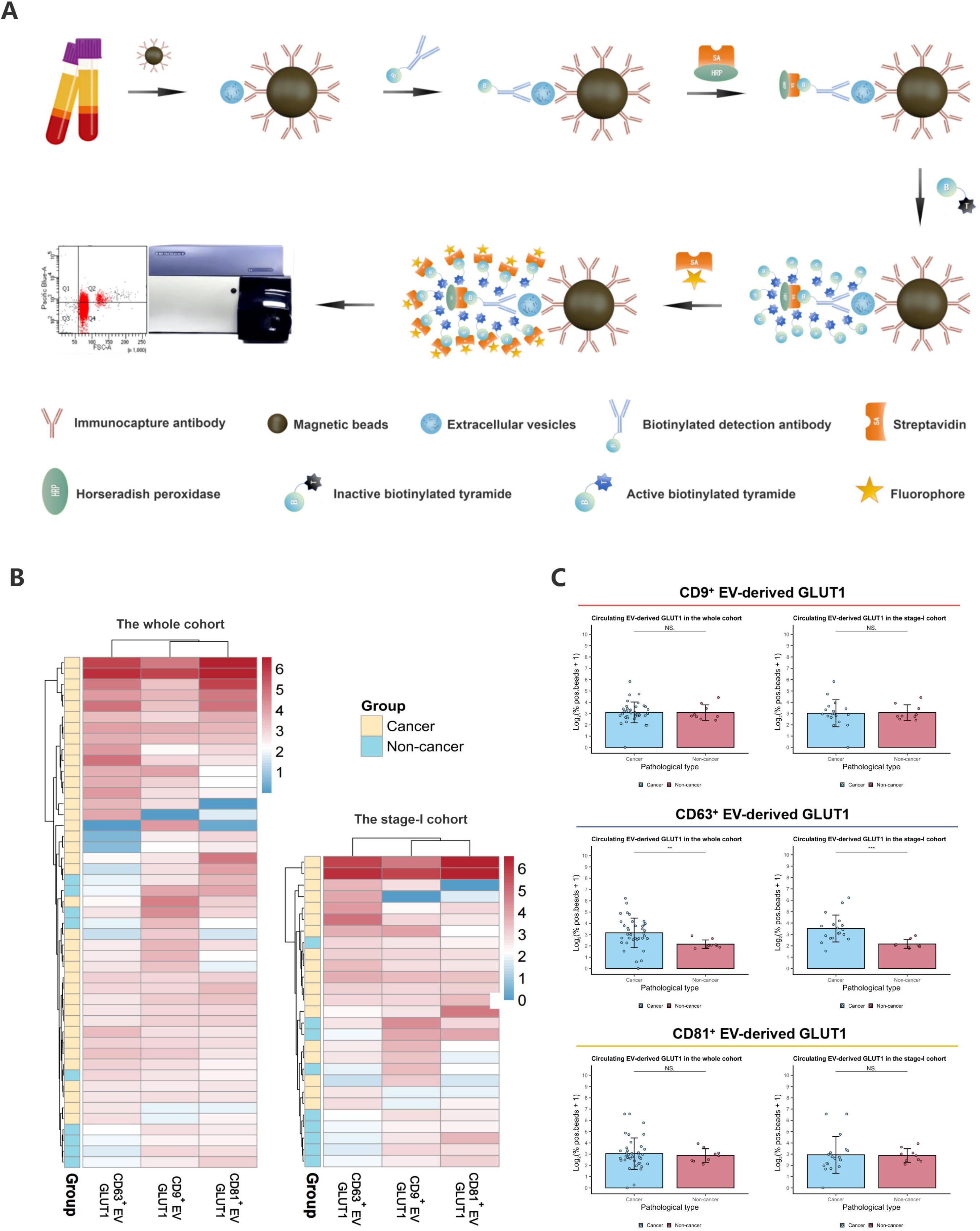
The modified DEST assay of circulating EV-derived GLUT1. (**A**) The schematic illustration of the modified DEST assay. (**B**) Hierarchical clustering heatmap of GLUT1 expression in circulating EVs, and (**C**) Box plots of GLUT1 expression in circulating EVs captured by CD9, CD63, and CD81. **Abbreviations**: DEST, digital extracellular vesicle screening technique; EV, extracellular vesicle; GLUT1, glucose transporter 1.

**Table 1.** The baseline characteristics of patients included in the study.

| Characteristics | Patients (%) |
| --- | --- |
|  | N = 47 |
| Age | 70.00 ± 9.71 |
| Gender |  |
| Male | 38 (80.85) |
| Female | 9 (19.15) |
| Smoking history |  |
| Yes | 38 (80.85) |
| No | 9 (19.15) |
| Emphysema |  |
| Yes | 24 (51.06) |
| No | 23 (48.94) |
| Malignant history |  |
| Yes | 8 (17.02) |
| No | 39 (82.98) |
| Pathological diagnosis |  |
| Cancer | 38 (80.85) |
| Non-cancer | 9 (19.15) |
| Subtype * |  |
| LUAD | 24 (51.06) |
| LUSC | 13 (27.66) |
| ASC | 1 (2.13) |
| Stage * |  |
| I | 18 (38.30) |
| II/III | 20 (42.55) |

\* Subtype and stage characteristics were collected from the 38 patients with
NSCLC, and were not applicable to the non-cancer patients. **Abbreviations:**
NSCLC, non-small cell lung cancer; LUAD, lung adenocarcinoma; LUSC, lung
squamous carcinoma; ASC, adenosquamous carcinoma lung cancer.

The well-established EV markers (CD9, CD63, and CD81) were utilized for immunocapture and subsequent analysis. Specifically, CD63^+^ EV-derived GLUT1 exhibited signal elevation in cancer patients, forming an independent cluster, whereas the CD9^+^ and CD81^+^ counterparts had similar expression patterns distinctly different from those of the CD63^+^ EVs (**Figure 5B**). Consistent clustering patterns were observed in the stage-I cohort. Moreover, the box plots confirmed significantly higher CD63^+^ EV-derived GLUT1 levels in the cancer samples, a finding that remained significant in the stage-Ⅰ subgroup (*P* < 0.05); in contrast, no significant differences in the CD9^+^ and CD81^+^ counterparts were observed between the cancer and non-cancer samples (**Figure 5C**). We observed no significant differences in the CD63⁺ EV-derived GLUT1 levels among the subgroups stratified by postoperative clinicopathological variables, including pathological stage, histological subtype, lymph node metastasis (LNM), recurrence, and epidermal growth factor receptor (EGFR) status (**Figure S3A-E**).

### 6. Diagnostic performance of circulating EV-derived GLUT1

The correlation heatmap revealed that pathological diagnosis was not strongly associated with preoperative clinical variables, except for CD63^+^ EV-derived GLUT1 expression (r = 0.70, **Figure 6A**). Notably, this correlation was substantially stronger in patients with stage-I NSCLC (r = 0.84, **Figure 6B**).

**Figure 6.**
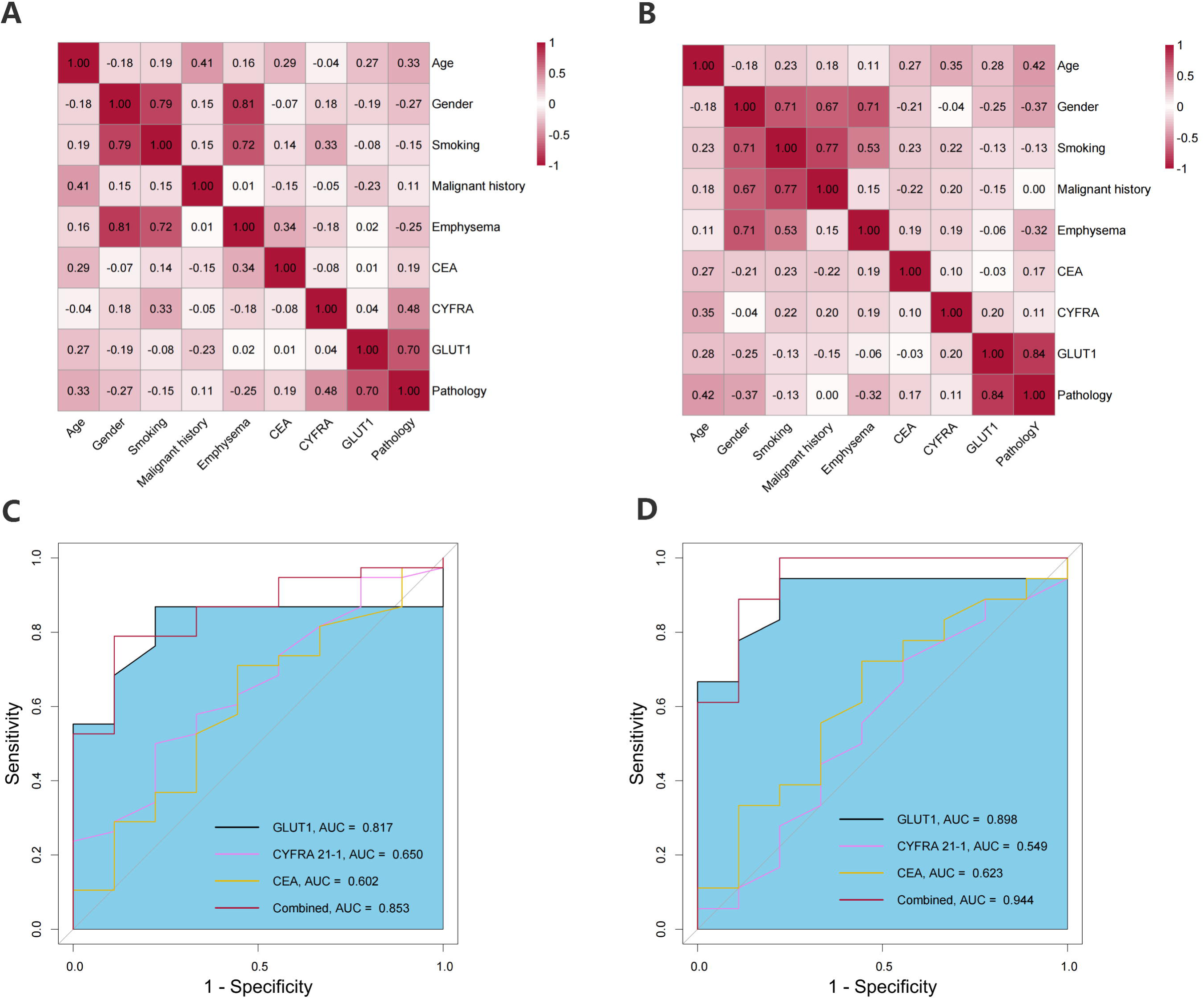
Diagnostic performance of circulating EV-derived GLUT1. (**A**) Correlation heatmap between GLUT1 expression and other clinicopathological characteristics in the whole cohort, and (**B**) in the stage-I cohort; pathology represents a binary variable (cancer and non-cancer). (**C**) ROC curve analysis comparing the diagnostic performance of CD63^+^ EV-derived GLUT1, CYFRA 21-1, CEA and the combination in the whole cohort, and (**D**) in the stage-I cohort. **Abbreviations**: EV, extracellular vesicle; ROC, receiver operating characteristic; AUC, area under the curve; GLUT1, glucose transporter 1; CEA, carcinoembryonic antigen; CYFRA 21-1, cytokeratin 19 fragment.

In the whole cohort, CD63^+^ EV-derived GLUT1 achieved an AUC of 0.817 in the ROC curve analysis, outperforming the conventional serum biomarkers CYFRA 21-1 (AUC = 0.650) and CEA (AUC = 0.602) in discriminating cancer from non-cancer samples. The diagnostic performance was further improved by the three-marker panel, which achieved an AUC of 0.853 (**Figure 6C**). Notably, even in the stage-Ⅰ cohort, CD63^+^ EV-derived GLUT1 maintained superior diagnostic performance (AUC = 0.898), and the combined three-marker panel reached the highest AUC of 0.944 (**Figure 6D**). ROC curves for CD9^+^ and CD81^+^ EV-derived GLUT1 were presented in **Figure S4A-D**. The Brier scores (0.116 and 0.095) and calibration curves indicated acceptable discrimination and calibration of the three-marker panel (CD63^+^ EV-derived GLUT1, CYFRA 21-1, and CEA) in the whole cohort and the stage-I group (**Table 2**, **Figure S4E-F**).

**Table 2.** The diagnostic performance of three-biomarker panel (CD63^+^ EV-derived GLUT1 + CEA + CYFRA 21-1).

| Models | AUC | Sensitivity | Specificity | Brier score |
| --- | --- | --- | --- | --- |
| **Whole cohort** | 0.853 (0.733-0.975) | 0.816 (0.526-0.947) | 0.889 (0.667-1.000) | 0.116 (0.064-0.173) |
| **Stage-I cohort** | 0.944 (0.852-1.000) | 1.000 (0.722-1.000) | 0.889 (0.667-1.000) | 0.095 (0.032-0.177) |

\* EV, extracellular vesicle; GLUT1, glucose transporter 1; CEA, carcinoembryonic
antigen; CYFRA 21-1, cytokeratin 19 fragment; AUC, area under the curve.

## Discussion

Despite the growing promise of EVs in oncology, the inherent heterogeneity and the low-abundance of EV cargoes have posed significant technical challenges, presenting a critical hurdle for moving the clinical translation forward [1, 6]. Based on an integrative analysis of tissue-derived EV proteomic data and public transcriptomic data, this study is the first to demonstrate the clinical utility of circulating CD63⁺ EV-derived GLUT1 as a biomarker for NSCLC. Its combination with conventional serum biomarkers enhanced diagnostic performance, particularly in early-stage cases.

Proteomic technologies have substantially enhanced our understanding of EV cargoes and their underlying biological mechanisms [4, 20]. For the discovery of minimally invasive EV-based LB biomarkers, previous studies conducted conventional proteomic analyses directly on circulating EVs [29–31]. Although some circulating complement components or immunoglobulins are genuinely associated with EVs, their high abundance enables coprecipitation during EV isolation, limiting the sensitivity of conventional proteomic approaches [20, 30–32]. Moreover, the high heterogeneity of circulating EV populations often dilutes cancer-specific signals [30, 31, 33, 34]. In this study, tissue-derived EVs, which had characteristics comparable to those of cell line-derived EVs, were subjected to proteomic analysis, which showed effective clustering of the protein profiles. Although extensive studies have focused on EVs isolated from body fluids or cell lines, the significant potential of tissue-derived EVs remains undiminished [35]. Beyond enabling the identification of highly cancer-specific biomarkers, tissue-derived EVs may better reflect the tumor microenvironment and provide more representative biological insights [35, 36]. Accordingly, a promising strategy for future investigations is to identify cancer-specific molecules from tissue-derived EVs and evaluate their potential as non-invasive circulating biomarkers [33, 36, 37].

The differentially expressed DEPs identified by the integrated proteomic and transcriptomic analyses in our study were enriched in the glucose metabolism- and hypoxia-associated pathways. Consequently, GLUT1 was prioritized for further analysis among the candidate biomarkers. GLUT1 is encoded by the SLC2A1 gene and functions as a facilitative membrane glucose transporter that is frequently overexpressed across various malignancies [38–40]. Upregulation of GLUT1 enhances glucose uptake and drives aerobic glycolysis (also known as the Warburg effect), positioning GLUT1 as a central mediator of cancer metabolic reprogramming [41–44]. The GLUT1-mediated metabolic shift primarily occurs via the HIF-1α and PI3K/AKT pathways, particularly under hypoxic conditions [45–49]. In recent years, accumulated evidence has demonstrated that EVs can transfer glycolytic phenotypes to recipient cells, thereby promoting oncogenesis by metabolic reprogramming [50, 51]. Primarily, EVs can encapsulate functional glycolytic proteins, such as GLUT1, which could be phagocytosed or fused with the plasma membrane of recipient cells, thereby enhancing glucose uptake and directly inducing Warburg-like metabolic reprogramming in recipient cells [52, 53]. In addition, EVs and their cargoes can modulate glycolytic pathways and activate intracellular signaling cascades in recipient cells, particularly promoting glucose reallocation and the establishment of an immunosuppressive premetastatic niche [54, 55]. Collectively, these findings demonstrate the critical role of GLUT1-associated EVs in metabolic reprogramming, highlighting their potential as therapeutic targets and biomarkers in oncology research. To date, EV-derived GLUT1 has not been reported in NSCLC, but non-invasive early detection will benefit from elucidating its oncological significance and clinical relevance.

The ultrasensitive TSA technique has become increasingly favored for detecting low-abundance proteins in various immunoassays [24, 25]. Yang et al. [26] introduced a DEST assay utilizing antibody pairs targeting different epitopes and integrating TSA to quantify EV-derived proteins from minimally processed plasma samples. The DEST assay achieved a diagnostic sensitivity up to 10^4^-fold greater than that of conventional ELISA and required < 4 hours to complete [26, 27]. We introduced a minor modification to the assay by immunocapturing EVs via tetraspanin binding, which markedly enhanced the capture efficiency and minimized the reliance on detection antibodies targeting specific epitopes. The results from our modified DEST assay demonstrated that CD63^+^ EV-derived GLUT1 had a clustering pattern distinctly different from the patterns of its CD9^+^ and CD81^+^ counterparts. Furthermore, CD63^+^ EV-derived GLUT1 exhibited significantly elevated signals in the cancer samples, suggesting that CD63-bearing EVs may correspond to a cancer-relevant subpopulation. Previous studies have suggested that EVs bearing only CD9 or CD81 predominantly originate from the plasma membrane (i.e., ectosomes), whereas those bearing CD63 along with one or both of the other tetraspanins are probably associated with endosome-derived exosomes, which are believed to have a more prominent role in intercellular communication [4, 56, 57]. Further studies into EV subpopulations are warranted to validate their molecular characteristics, ultimately guiding the design of optimal immunocapture strategies for circulating EV-based LB in clinical practice.

Several serum biomarkers, including CEA and CYFRA 21-1, have been widely used for detection and risk stratification in NSCLC [58, 59]. Integrating multiple biomarkers into risk prediction models, such as the four-marker protein panel (4MP) model or lung cancer biomarker panel (LCBP) model, has enhanced sensitivity and specificity compared to a single-marker approach [58–60]. However, the limited biological specificity and heterogeneous expression of conventional biomarkers remain significant challenges for the early detection of NSCLC. In this study, circulating EV-derived GLUT1 demonstrated diagnostic performance superior to that of the conventional biomarkers, CEA and CYFRA 21-1, for discriminating between cancer from non-caner samples. The combined panel incorporating CD63^+^ EV-derived GLUT1, CYFRA 21-1, and CEA posed enhanced the discriminative capacity relative to that of either alone. Notably, this superiority could be extended to early-stage patients. From a clinical applicability perspective, validation using the DEST assay in larger and external cohorts is necessary, preferably by integrating classic models to establish a comprehensive diagnostic panel, which could potentially improve non-invasive early detection and stratification of NSCLC.

Several limitations of this study should be acknowledged when interpreting the results. First, the sample size was small and obtained from a single-center retrospective cohort, potentially limiting the statistical robustness and generalizability of our findings. Second, although signal amplification enhanced detection sensitivity, it may have introduced background noise; reliance on specific antibodies could also lead to variability depending on antibody specificity and epitope availability. Combining multiple biomarkers may improve the specificity of cancer-derived EV protein detection. Finally, our study concentrated on the clinical performance of circulating EV-derived GLUT1 without providing mechanistic insight into the oncogenic pathways. Therefore, further research is warranted to investigate its biogenesis, release, uptake, and downstream effects.

## Conclusion

The findings of this study support the use of a promising biomarker discovery strategy that identifies candidate markers from tissue-derived EVs and validate their diagnostic performance in circulation. This approach could enhance the cancer specificity of EV-based biomarkers in early detection. By performing a modified DEST assay, we identified circulating EV-derived GLUT1 as a novel biomarker for early-stage NSCLC, with improved discriminative capacity when combined with conventional serum biomarkers. These findings underscore the feasibility of translating tissue-derived EV proteomic discoveries into LB-based early detection strategies.

## Data Availability

All data produced in the present study are available upon reasonable request to the authors.

## Acknowledgments

The authors wish to acknowledge the Division for Medical Research Engineering, Nagoya University Graduate School of Medicine for the usage of NanoSight NS300 system, Optima XE-100, and BD FACS Canto™ II Flow Cytometer, and we also thank Dr. Itakura (Technical Center, Nagoya University) for his technical support and data acquisition. In addition, we gratefully acknowledge Enago company (www.enago.jp) for English language editing.

## Conflict of interest

There is no conflict to declare.

## Funding

This work was supported by JSPS KAKENHI (Grant number: 22K16566 and 24K12009), and the Hori Sciences and Arts Foundation and Nagoya University Hospital Funding for Clinical Research.

## Author contributions

Conception and design, H.H. and T.K.; Provision of study materials, H.H., Y.I., Y.N., H.W., H.T., T.R., Y.K., K.N., Y.K., and H.U.; Collection and assembly of data, H.H., T.K., Y.A., A.Y., M.K., and E. A-I.; Data analysis and interpretation, H.H., T.K., S.N., T.M., A.T., A.Y., and T. C-Y.; Manuscript writing: All authors; Administrative support, T.K. and T. C-Y. Final approval of manuscript: All authors.

## Data sharing statement

The data that support the findings of this study are available from the corresponding authors upon reasonable request.

## Supplementary materials

**Table S1. Experimental protocols of the modified DEST assay.**

* Beads are collected using a 96-well plate magnet in each washing step. Each incubation is performed on a plate shaker to prevent the beads deposition. Washing times could be adjusted according to experimental requirement. **Abbreviations**: DEST, digital extracellular vesicle screening technique; BSA, bovine serum albumin; PBS, phosphate buffered saline; HRP, horseradish peroxidase; FACS, fluorescence-activated cell sorting.

**Table S2.** List of reagents used in the modified DEST assay.

* **Abbreviations**: DEST, digital extracellular vesicle screening technique; BSA, bovine serum albumin; PBS, phosphate buffered saline; GLUT1, glucose transporter 1; mAb, monoclonal antibody; HRP, horseradish peroxidase; TSA, tyramide signal amplification; DMSO, dimethyl sulfoxide.

**Figure S1. Identification and functional annotation of the DEGs from the TCGA dataset.** (**A**) Volcano plot visualizing the distribution, (**B**) Hierarchical clustering heatmap (**C**) GO enrichment analysis, and (**D**) KEGG pathway enrichment analysis of DEGs from the TCGA dataset. (**E**) GO enrichment analysis, and (**F**) KEGG pathway enrichment analysis of the 18 cancer-specific DEGs candidate target biomarkers. **Abbreviations**: DEPs, differentially expressed genes; TCGA, The Cancer Genome Atlas; GO, Gene Ontology; KEGG, Kyoto Encyclopedia of Genes and Genomes.

**Figure S2. The gene expression and diagnostic performance of the other seven candidate genes.** (**A**) Box plots of differential expression, and (**B**) ROC curve analyses of the other seven candidate target biomarkers between tumor and normal tissues from the TCGA dataset. **Abbreviations**: TCGA, The Cancer Genome Atlas; ROC, receiver operating characteristic; AUC, area under the curve; CI, confidence interval.

* All analyses in panels A and B were performed on 106 paired tumor-normal tissue samples from the TCGA dataset.

**Figure S3. Circulating EV-derived GLUT1 expression across clinicopathological characteristics.** (**A**) Box plots illustrating CD63⁺ EV-derived GLUT1 expression stratified by pathological stage, (**B**) histological subtype, (**C**) LNM, (**D**) recurrence, (**E**) EGFR status. **Abbreviations**: EV, extracellular vesicle; vesicle; GLUT1, glucose transporter 1; LNM, lymph node metastasis; EGFR, epidermal growth factor receptor.

**Figure S4. ROC curve analyses and calibration curves and for circulating EV-derived GLUT1.** (**A**) ROC curve analysis comparing the diagnostic performance of CD9^+^ EV-derived GLUT1, CYFRA 21-1, CEA and the combination in the whole cohort, and (**B**) in the stage-I cohort. (**C**) ROC curve analysis comparing the diagnostic performance of CD81^+^ EV-derived GLUT1, CYFRA 21-1, CEA and the combination in the whole cohort, and (**D**) in the stage-I cohort. (**E**) Calibration curves for the three-marker panel (CD63^+^ EV-derived GLUT1, CYFRA 21-1 and CEA) in the whole cohort, and (**F**) in the stage-I cohort. **Abbreviations**: ROC, receiver operating characteristic; AUC, area under the curve; EV, extracellular vesicle; GLM, generalized linear model; GLUT1, glucose transporter 1; CEA, carcinoembryonic antigen; CYFRA 21-1, cytokeratin 19 fragment.

## Notes

### Competing Interest Statement

The authors have declared no competing interest.

### Author Declarations

Ethics committee/IRB of the Institutional Review Board of Nagoya University Hospital gave ethical approval for this work (No.2022-0107).

