## Supplementary figures and images for "An intratumorally discovered extracellular vesicle-derived biomarker enables ultrasensitive digital quantification in plasma for early detection of non-small cell lung cancer"

### Figure S1.JPEG

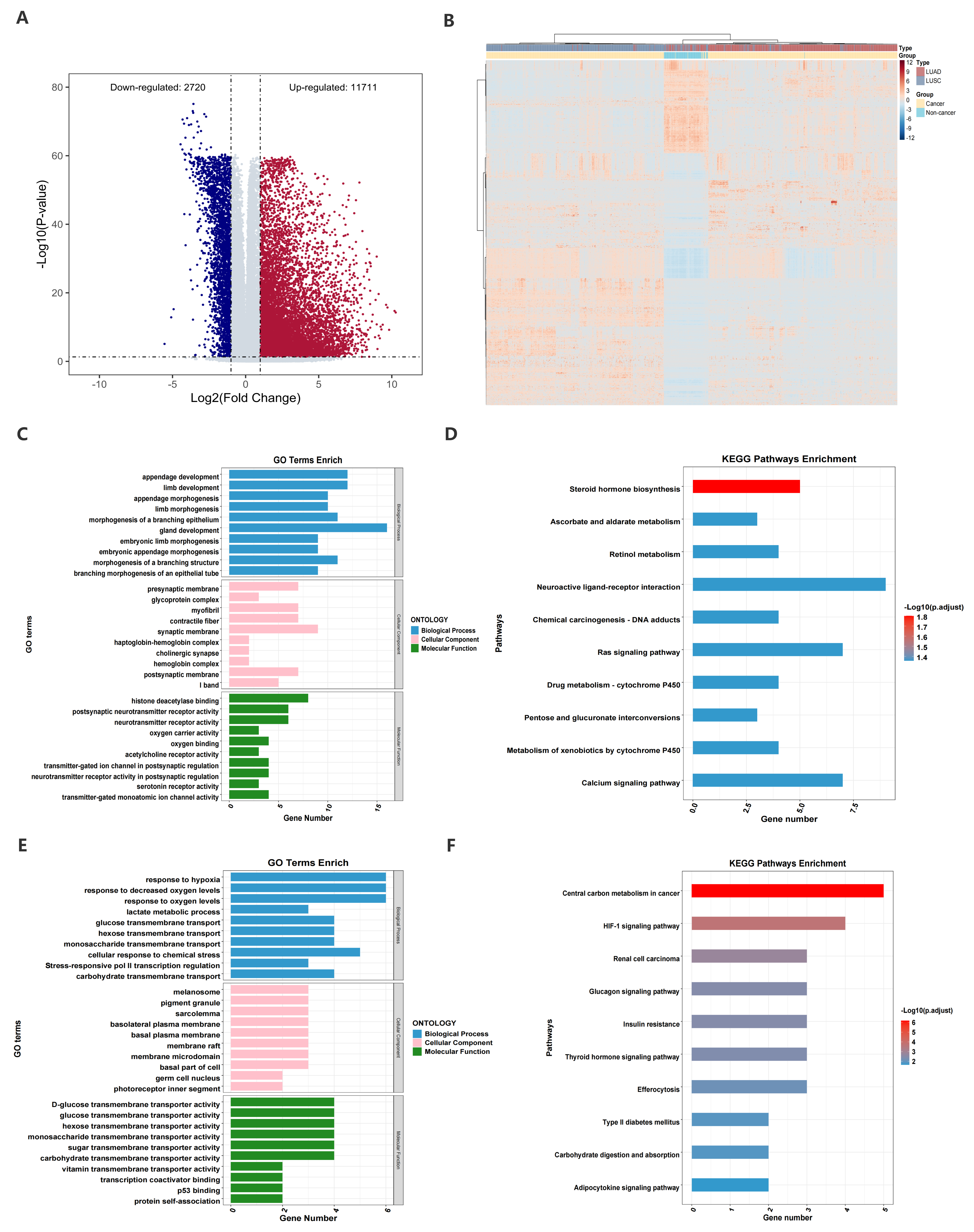

### Figure S2.JPEG

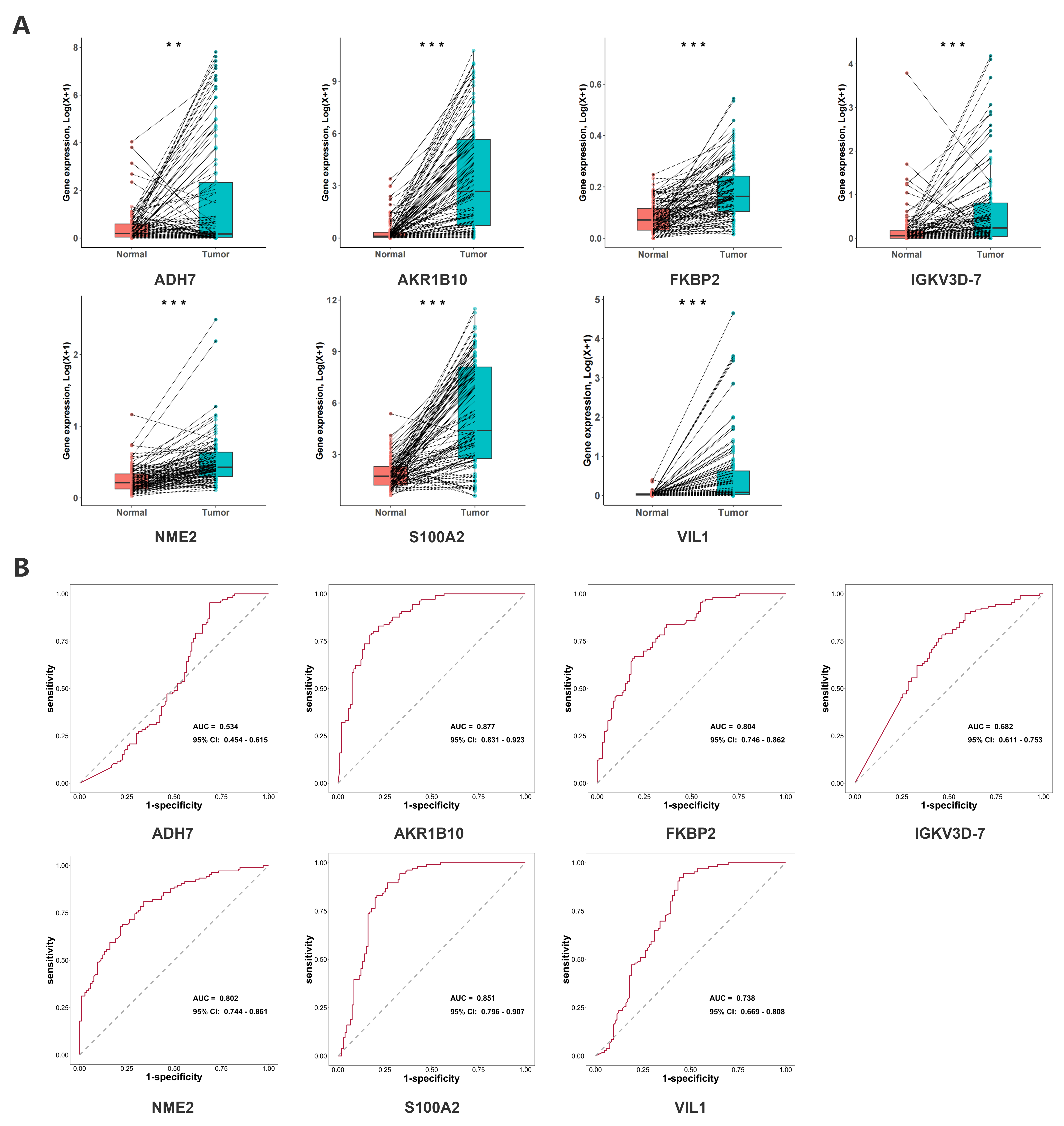

### Figure S3.JPEG

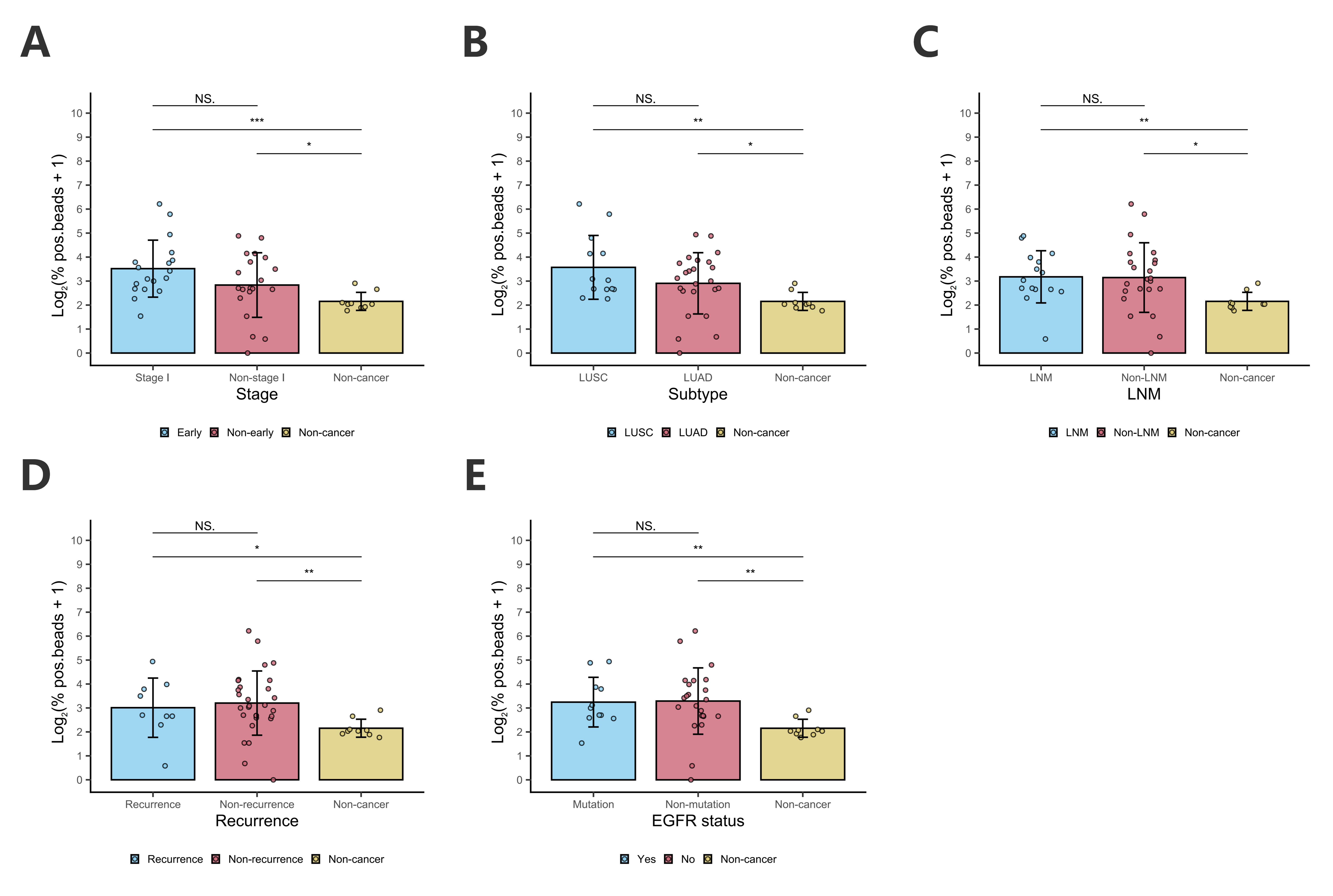

### Figure S4.JPEG

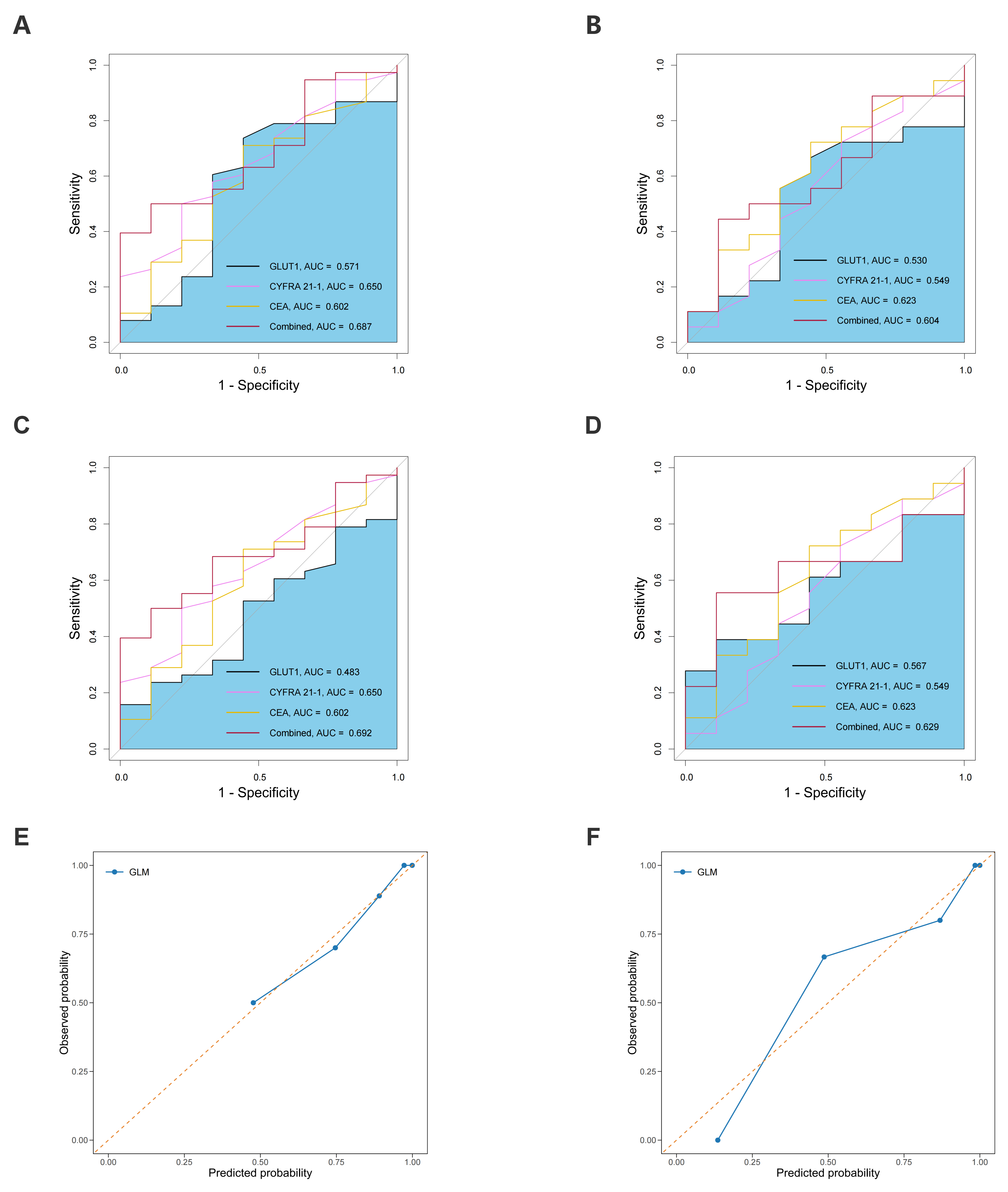
