## Supplemental Table 1 for "An intratumorally discovered extracellular vesicle-derived biomarker enables ultrasensitive digital quantification in plasma for early detection of non-small cell lung cancer": Table S1.docx

**Table S1. Experimental protocols of the modified DEST assay.**

| **Step** | **Reagent / Procedure** | **Time (min)** | **Volume (μl)** | **Concentration** | **Buffer** |
| --- | --- | --- | --- | --- | --- |
| 1 | Magnetic beads | 30 | 100 | 2 μl/well | 2% BSA |
| 2 | Washing × 4 |  | 100 |  | PBS + 0.1% Tween-20 |
| 3 | Plasma sample | 60 | 100 | 5 μl | 2% BSA |
| 4 | Washing × 4 |  | 100 |  | PBS + 0.1% Tween-20 |
| 5 | Detection antibody | 60 | 50 | 0.5 μg/ml | 2% BSA |
| 6 | Washing × 4 |  | 100 |  | PBS + 0.1% Tween-20 |
| 7 | Streptavidin-HRP | 30 | 100 | 1:200 | 2% BSA |
| 8 | Washing × 4 |  | 100 |  | PBS + 0.1% Tween-20 |
| 9 | Biotinylated tyramide | 10 | 100 | 1:50 | 1X plus amplification diluent |
| 10 | Washing × 4 |  | 100 |  | PBS + 0.1% Tween-20 |
| 11 | Fluorescence-labeled streptavidin | 30 | 50 | 1:1000 | 2% BSA |
| 12 | Washing × 4 |  | 100 |  | PBS + 0.1% Tween-20 |
| 13 | FACS test |  | 250 |  |  |

^*^ Beads are collected using a 96-well plate magnet in each washing step. Each incubation is performed on a plate shaker to prevent the beads deposition. Washing times could be adjusted according to experimental requirement. **Abbreviations**: DEST, digital extracellular vesicle screening technique; BSA, bovine serum albumin; PBS, phosphate buffered saline; HRP, horseradish peroxidase; FACS, fluorescence-activated cell sorting.
