## Supplemental Table 1 for "An intratumorally discovered extracellular vesicle-derived biomarker enables ultrasensitive digital quantification in plasma for early detection of non-small cell lung cancer": Table S2.docx

**Table S2. List of reagents used in the modified DEST assay.**

| **Product name** | **Company** | **Catalog number** | **Specification** | **Concentration used** |
| --- | --- | --- | --- | --- |
| CD9 (D8O1A) Rabbit mAb | Cell Signaling | 13174S | 84 μg/ml, 100 μl | 84 μg/ml, 100 μl |
| CD63 (D4I1X) Rabbit mAb | Cell Signaling | 55051S | 11 μg/ml, 100 μl | 11 μg/ml, 100 μl |
| CD81 (D3N2D) Rabbit mAb | Cell Signaling | 56039S | 50 μg/ml, 100 μl | 50 μg/ml, 100 μl |
| Dynabeads™ Antibody Coupling Kit | Thermo Fisher | 14311D | 60 mg | 5 mg/500 μl |
| BSA | Sigma-Aldrich | A7030 | 100 g | 2% |
| PBS | Thermo Fisher | 10010-023 | 1X/500 ml | - |
| Tween-20 | Sigma-Aldrich | P1379 | 100ml | 0.1% |
| Zeba™ Spin Desalting Columns | Thermo Fisher | 89882 | 0.5 ml | - |
| EZ-Link™ Sulfo-NHS-LC-Biotin | Thermo Fisher | A39257 | 1 mg | 1 mg/180 μl |
| Anti-Glucose Transporter GLUT1 antibody | Abcam | ab115730 | 0.179 mg/ml, 100 μl | 0.5 μg/ml |
| Streptavidin-HRP | R&D Systems | DY998 | 1 ml, 1:200 | 1:200 |
| TSA Plus Biotin | Akoya Biosciences | NEL749A001KT | Dissolve in 300 μl DMSO | 1:50 |
| Brilliant violet 421 streptavidin | BioLegend | 405225 | 0.5 mg/ml, 200 μl | 1:1000 |

^*^ **Abbreviations**: DEST, digital extracellular vesicle screening technique; BSA, bovine serum albumin; PBS, phosphate buffered saline; GLUT1, glucose transporter 1; mAb, monoclonal antibody; HRP, horseradish peroxidase; TSA, tyramide signal amplification; DMSO, dimethyl sulfoxide.
